# Closed-loop neurostimulation for epilepsy leads to improved outcomes when stimulation episodes are delivered during periods with less epileptiform activity

**DOI:** 10.1101/2022.11.28.22282784

**Authors:** Daria Nesterovich Anderson, Chantel M. Charlebois, Elliot H. Smith, Tyler S. Davis, Angela Y. Peters, Blake J. Newman, Amir M. Arain, Karen S. Wilcox, Christopher R. Butson, John D. Rolston

## Abstract

In patients with drug-resistant epilepsy, electrical stimulation of the brain in response to epileptiform activity can make seizures less frequent and debilitating. When effective, this therapy, known as closed-loop responsive neurostimulation (RNS), produces long-lasting changes in brain dynamics that correlate with clinical outcomes. Since periods with frequent epileptiform activity are less conducive to neuroplasticity, we hypothesize that stimulation timing, specifically stimulation during brain states with less epileptiform activity, is critical in driving long-term changes that restore healthy brain networks. To test this, we quantified stimulation episodes during low- and high-risk epochs—that is, stimulation during periods with a low or high risk of generating seizures and less or more epileptiform activity—in a cohort of 40 patients treated with RNS. Patients were categorized into three groups: super responders (>90% reduction, n=10), intermediate responders (≥ 50% reduction and ≤ 90% reduction), n=19, and poor responders (<50% reduction, n=11). As hypothesized, in this retrospective study, seizure reduction (median 64.6% reduction at last follow-up) was correlated with more frequent stimulation during low-risk periods compared with high-risk periods. Additionally, stimulation events were more likely to be phase-locked to prolonged episodes of abnormal activity for intermediate and poor responders when compared to super responders, consistent with the hypothesis that improved outcomes are driven by stimulation during low-risk states. These results suggest that stimulation during low-risk periods may more readily induce plasticity that, in turn, facilitates network changes leading to long-term seizure reduction.

**One Sentence Summary:** Increased stimulation during periods of reduced seizure risk corresponds with improved therapy in responsive neurostimulation for epilepsy.

## INTRODUCTION

Closed-loop responsive neurostimulation (RNS) is an FDA-approved therapy for treatment-resistant epilepsy, used when resection or lesioning is not an option. While true seizure freedom is rare, seizure reduction is common with RNS and improves over time. The initial clinical trial for the RNS system reported a 53% median decrease in seizure frequency and 12.9% seizure free period of least one year, at two years (Morrell, 2011; Bergey *et al*., 2015), while 9-year outcomes boast 75% median decrease in seizure frequency and 18.4% seizure freedom rates for at least a one-year period (Nair *et al*., 2020). Responsive neurostimulation not only has an acute effect via a reduction in spectral power post-stimulation (Rønborg *et al*., 2021), but continued therapy leads to long-term improvements (Nair *et al*., 2020).

Improved clinical outcomes have correlated with various features of neural signals recorded on RNS. For instance, previously reported “indirect effects” of stimulation, such as spontaneous ictal inhibition or altered frequency dynamics during seizures, correlated with better clinical outcomes while acute, direct effects of stimulation did not correlate with outcomes (Kokkinos *et al*., 2019). Furthermore a cohort of super-responder neuromodulation patients (>90% improvement) experienced functional network reorganization while poor responders (< 50% improvement) did not experiences these network changes (Khambhati *et al*., 2021). Taken together, these results suggest that network reorganization might drive improved clinical outcomes. Determining the factors that enable plastic, functional changes over time from those that do not will be critical for understanding and improving response to neuromodulation.

Based on evidence that synapses may already be saturated after the seizure event, thereby impairing long-term plasticity after a seizure (Beck *et al*., 1996; Abegg *et al*., 2004; Naik *et al*., 2021), it is unlikely that long-lasting network changes occur from stimulation occurring during seizures. While stimulation during a seizure might act through acute desynchronization to arrest a seizure (Lesser *et al*., 1999), we hypothesized that stimulation occurring during periods of relative ictal quiescence, leads to long-lasting functional network changes that are correlated with improved outcomes.

To test this hypothesis, we divided patient stimulation into periods with higher risk (high-risk epoch) or lower risk (low-risk epoch) of generating seizures and correlated the respective stimulation to clinical outcomes. Determining whether stimulation is in a high- or low-risk epoch may be estimated based on stimulation counts and repetitive detections, known as long episodes, collected from each patients’ intracranial device (Chiang *et al*., 2021). We found early biomarkers of RNS efficacy, namely that stimulation in low-risk epochs promoted improved clinical outcomes.

## RESULTS

Outcomes and demographic variables were gathered for 40 patients implanted with responsive neurostimulation devices (Fig 1A). Clinical outcomes were collected after patients had devices implanted for a median time of 860 days (LQ: 522 days, UQ: 1324 days). Patients experienced a significant reduction in seizures with responsive neuromodulation therapy (Fig 1B; one sample *t*-test; p < 0.0001). Median seizure reduction rates at follow-up were 64.6% (Fig 1B; LQ: 37.1%, UQ: 90.8%). Using this patient cohort, we calculated how much time and how many stimulation episodes patients experienced according to methods described in Figure 1D, and defined periods with below average epileptiform activity to be “low risk”, similar in concept to Chiang *et al*., 2021.

**Fig. 1.**
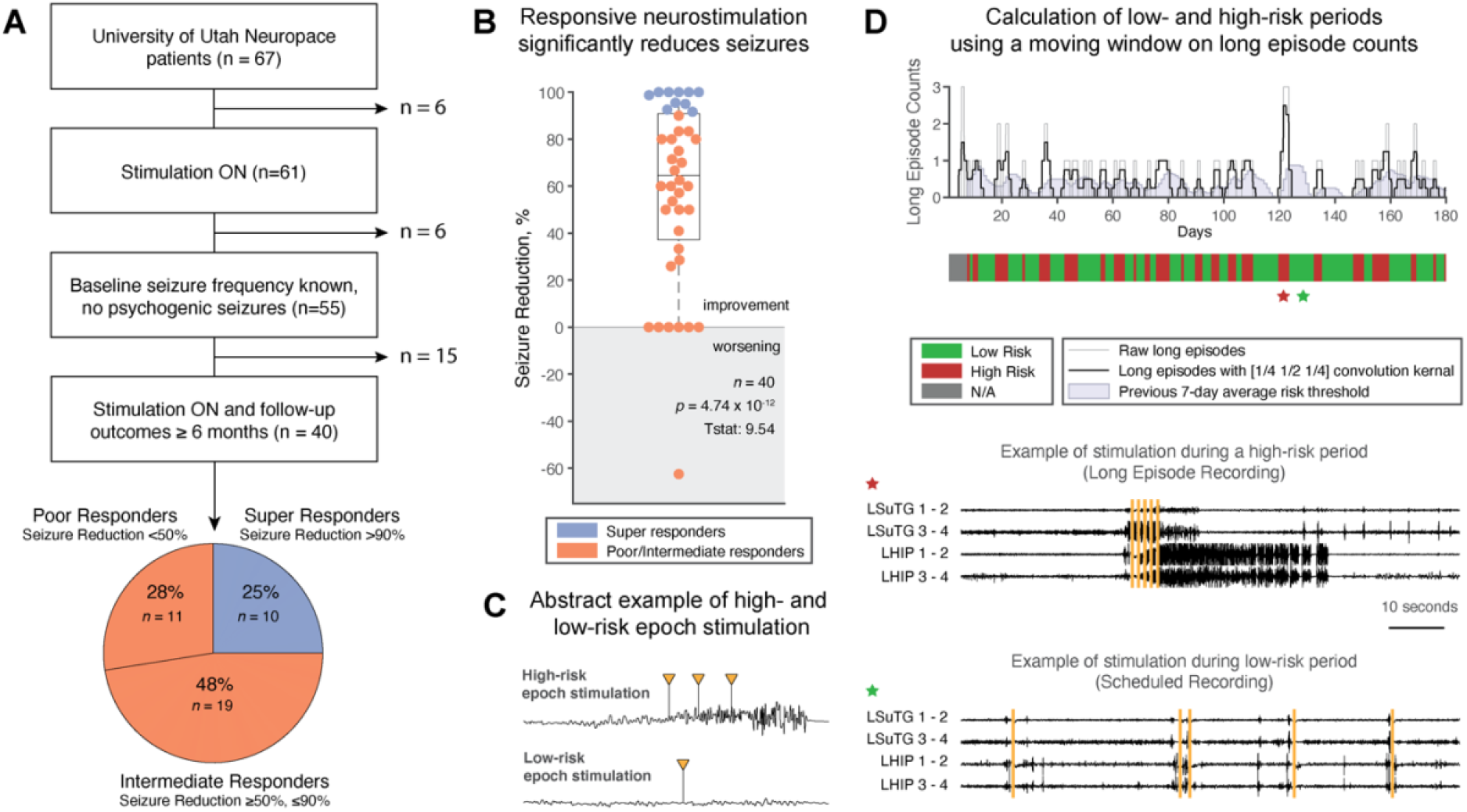
Patient cohort summary and determination of high- and low-risk epochs. (**A**) Summary of patients incorporated into the study. 40 patients were included in the study: 10 super responders, 19 intermediate responders, and 11 poor responders. (**B**) The RNS System is effective at reducing seizure frequency (one sample *t*-test; p < 0.0001) with a median seizure reduction of 64.6% (LQ: 37.1%, UQ: 90.8%). (**C**) We provide a theoretical example of a high-risk epoch stimulation event we hypothesize will not lead to long-term network changes versus low-risk epoch stimulation that we hypothesize leads to long-term network changes. (**D**) Demonstration of risk period calculation for a 6-month period in an example patient. Long episode counts are extracted from the RNS System, and convolved with a [1/4, 1/2, 1/4] day kernel window to calculate risk amplitude. A day is considered high risk if the calculated daily risk is greater than the previous 7-day average of risk amplitudes. Example ECoG traces from periods of high risk and low risk are shown to parallel our hypothesis in Figure 1C. Yellow bars indicate deployed stimulation.

### Failure to predict seizure reduction from standard metrics

We tested the hypothesis that an increased number of stimulation episodes delivered during periods of reduced epileptic activity is predictive of improved patient outcomes (Fig 1C). The earlier it may be possible to determine predictors of patient outcomes, the more beneficial for patients (Fig 2A). Baseline seizure frequency, known prior to surgical intervention, was not able to predict subsequent seizure reduction (Fig 2B). Additionally, patient age, age of epilepsy onset, and epilepsy durations were not predictive of seizure reduction (Fig S1). Prior to stimulation being enabled, there was a median period of 36.5 days (LQ: 21 days, UQ 61 days) where the RNS System was implanted and detection protocols were similar across all patients, but stimulation was not yet turned on. We found that long episode counts, or prolonged and repeated detections, measured during this baseline period significantly correlated with monthly baseline seizure rate (Fig 2C; Pearson’s correlation, *p* = 0.011), signifying that long episodes aptly quantify seizure burden. However, the number of baseline long episodes were not significantly correlated with seizure reduction, and therefore were not able to predict outcomes early in therapy (Fig 2D; Pearson’s correlation, *p* = 0.30). Further, we neither saw a significant correlation in the number of baseline detections, linked to interictal epileptiform discharges (Baud *et al*., 2018; Gregg *et al*., 2021; Leguia *et al*., 2021), with monthly baseline seizure rates (Fig 2E; Pearson’s correlation, *p* = 0.22), nor a significant correlation between baseline detections with clinical outcomes (Fig 2F; Pearson’s correlation, *p* = 0.61). The number of stimulation episodes was initially similar between super-responders and intermediate/poor responders during the first 90 days (Fig. 2G), but the intermediate/poor responder groups experienced a steady increase in stimulation counts over the duration of their therapy (Fig 2H; Pearson’s correlation, *p*<0.0001), likely due to intentional changes in detection parameters by the epileptologists.

**Fig. 2.**
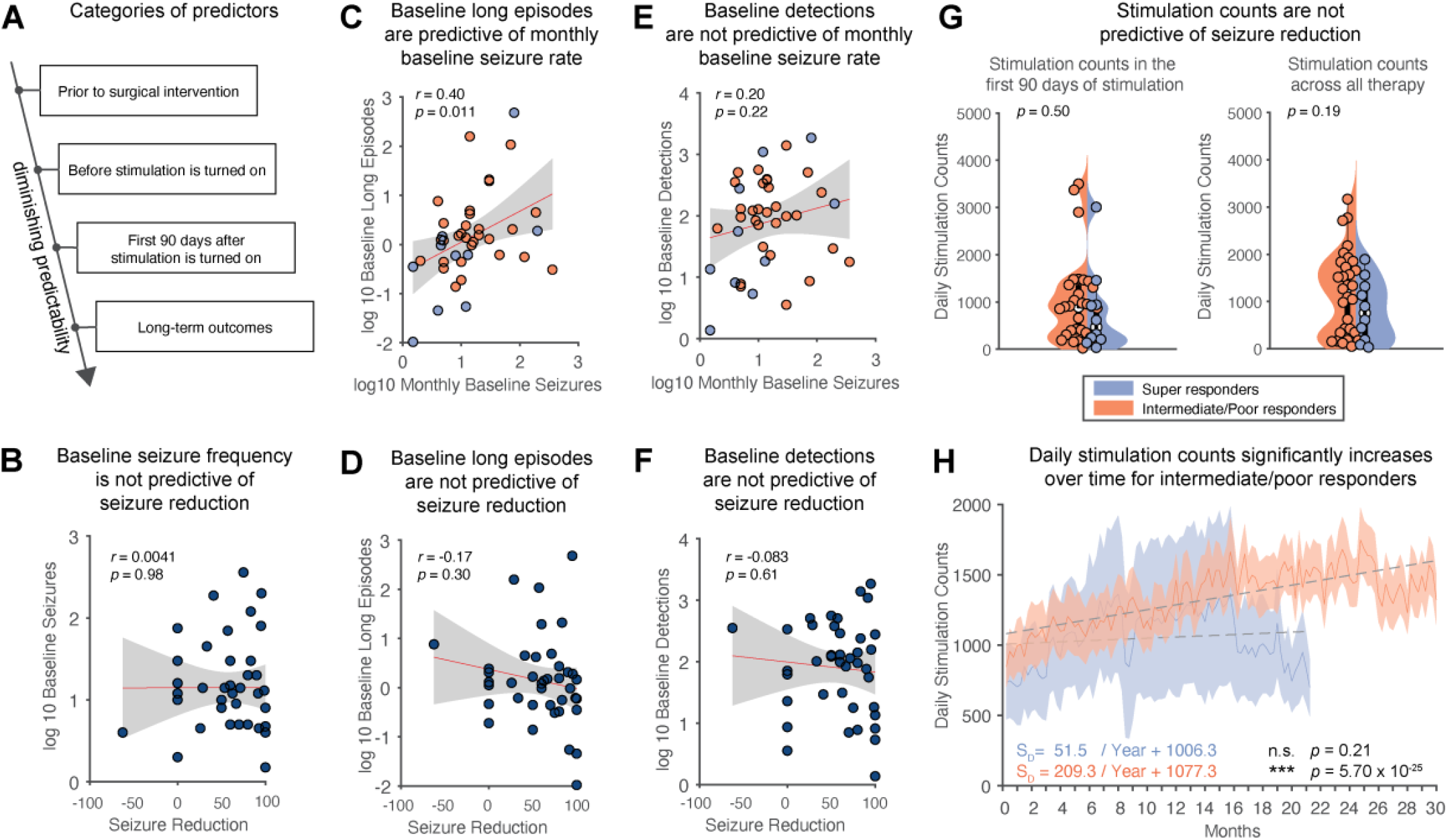
Seizure reduction is not significantly correlated with baseline seizure frequency, baseline detections, or stimulation counts. (**A**) Factors that can predict outcomes to neuromodulation have the most benefit if utilized earlier in therapy. (**B**) There is no correlation between baseline seizure rate and outcome (Pearson’s correlation; *p* = 0.98). (**C**) Prior to stimulation onset, in the baseline period, long episode counts are significantly correlated with monthly baseline seizures, indicating that long episodes correspond to baseline seizure rates. (Pearson’s correlation; *p* = 0.011). (**D**) However, these baseline long episodes are not significantly correlated with seizure reduction. (Pearson’s correlation; *p* = 0.30). (**E**) Baseline detections are additionally not correlated with monthly baseline seizures (Pearson’s correlation; *p* = 0.22). (**F**) Similarly, these baseline detections are not significantly correlated with seizure reduction (Pearson’s correlation; *p* = 0.62). (**G**) Daily stimulation counts between super responders and intermediate/poor responders are not significantly different either from the first 90 days of stimulation or over all stimulation time (two-sample *t*-test, *p* = 0.50; two-sample *t*-test, *p* = 0.19). (**H**) Tracking stimulation counts over time, intermediate/poor responders have an increase in stimulation over time (Pearson’s correlation; *p* < 0.0001).

### Increased stimulation in low-risk periods predicts outcomes

We calculated time spent in low-risk periods for each patient for the first 90 days of stimulation according to methods described in Fig 1D. From the onset of therapy, an increased time spent in low-risk states was correlated and predictive of seizure reduction (Fig 3A; Pearson’s correlation; *p* = 1.88 × 10^−3^, Leave-one-out cross validation: *p* = 0.011). Similarly, an increase in the ratio of stimulation episodes occurring during low-risk periods was correlated and predictive of seizure reduction (Fig 3B; Pearson’s correlation; *p* = 3.13 × 10^−3^, Leave-one-out cross validation: *p* = 0.019). Time spent in low-risk states significantly increased over time for intermediate/poor responders (Pearson’s correlation, *p* = 4.28 × 10^−4^), which may correspond to long-term improvements reported in prior patient cohorts along multi-year time scales (Nair *et al*., 2020). However, even after nearly 2.5 years of stimulation, intermediate/poor responders still spent less time in low-risk states on average than super responders at any time point.

**Fig. 3.**
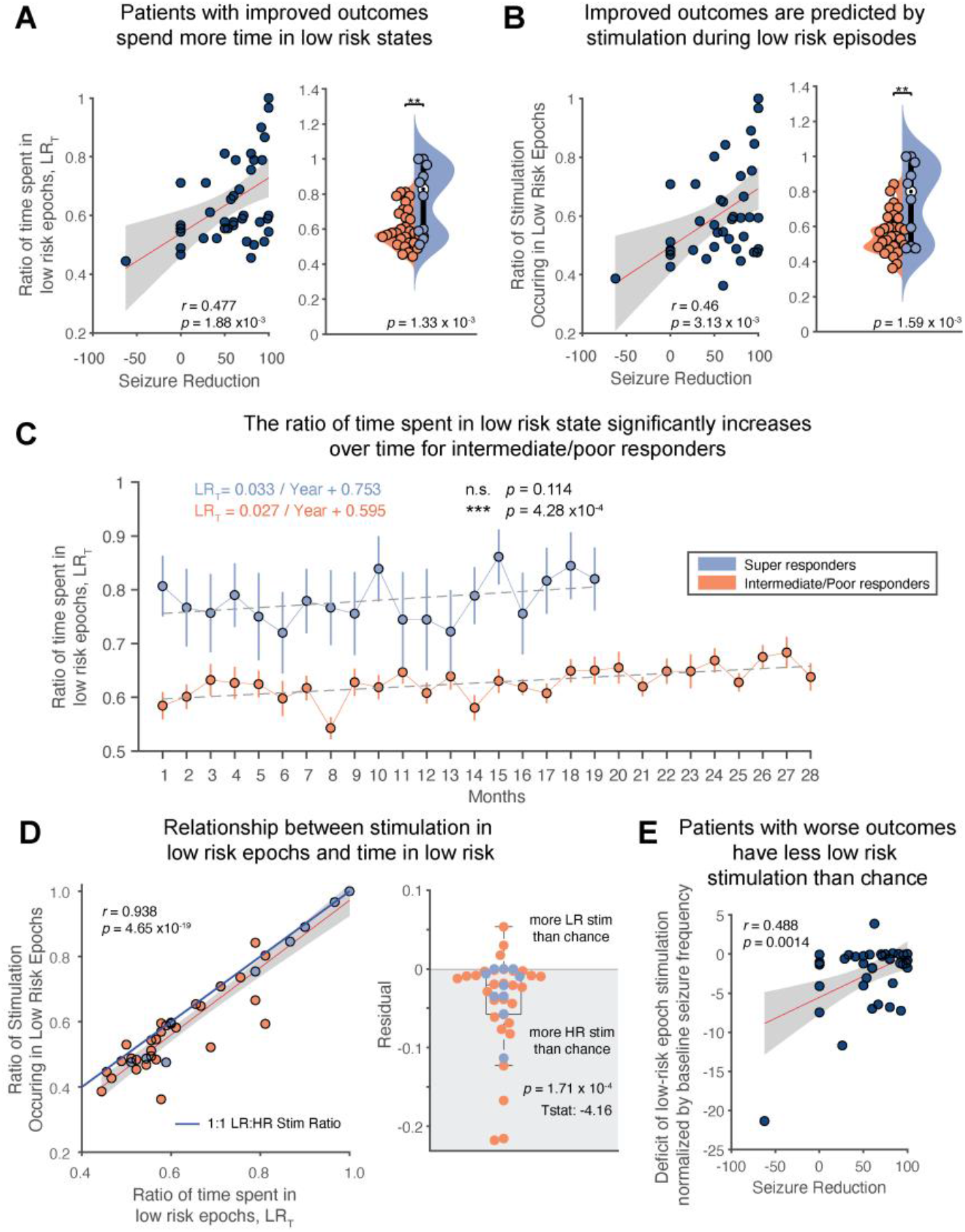
Increased time spent in low-risk epochs and more stimulation occurring during low-risk epochs is predictive of clinical outcomes. (**A**) Patients with greater seizure reduction spend a greater amount of time in low-risk states (Pearson’s correlation; *p* = 1.88 × 10^−3^). This analysis holds up to leave one out cross-validation (Pearson’s correlation; *p* = 0.011). Super responder patients spend significantly more time in low-risk states than high-risk states in the first 3 months of stimulation (two-sample *t*-test, *p* = 1.33 × 10^−3^). (**B**) Patients with greater seizure reduction also experience more stimulation during low-risk epochs (Pearson’s correlation; *p* = 3.13 × 10^−3^). This analysis also holds up to leave one out cross-validation (Pearson’s correlation; *p* = 0.019). Super responder patients receive significantly more stimulation during low-risk states than high-risk states in the first 3 months of stimulation (two-sample *t*-test, *p* = 1.59 × 10^−3^). (**C**) Tracking time in low-risk states over the course of stimulation, we find that time in low-risk states increases significantly over time for intermediate/poor-responders (Pearson’s correlation: *p*=4.28 × 10^−4^). Despite this, after nearly 2.5 years, intermediate/poor responders have not yet reached the ratio of time in low risk that super responders began at. (**D**) Panels **A** and **B** are highly correlated (Pearson’s correlation; p = 4.65 × 10^−19^). Compared to the solid blue line, which indicates a perfect balance between high-risk and low-risk stimulation, most RNS System patients have more high-risk stimulation than chance (one sample *t*-test; *p* = 1.71 × 10^−4^). (**E**) Having more high-risk stimulation episodes per seizure than chance correlates with poor clinical outcomes (Pearson’s correlation; *p* = 1.4 × 10^−3^).

We next asked how independent these two measures—time in low-risk states and stimulation during low-risk states—were. That is, were patients who initially spend more time in low-risk states fated to do better or does stimulation during these low-risk states confer an added benefit? To better dissect this relationship, we correlated time in low-risk states (Fig 3A) with low-risk stimulation (Fig 3B). While these two metrics were highly correlated (Pearson’s correlation; p = 4.65 × 10^−19^), nearly all patients deviated from what would be expected if stimulation were delivered uniformly or by chance. As expected from a closed-loop device triggering stimulation upon the detection of epileptiform activity, most patients’ stimulation episodes were biased to high-risk periods (Fig 3D; one sample *t*-test; *p* = 1.71 × 10^−4^). Graphically, the solid blue line in Figure 3D indicates a theoretical situation in which high-risk and low-risk stimulation episodes occur in identical proportions to the time spent in high- and low-risk states, a relationship that would be followed during open-loop stimulation (as in deep brain stimulation). Most patients treated with the RNS System fell below this line.

We next asked whether deviation from this “unbiased” allocation of stimulation was correlated with seizure reduction. Indeed, having more high-risk stimulation episodes per seizure was correlated with diminished seizure reduction (Fig 3E; Pearson’s correlation; *p* = 1.4 × 10^−3^), supporting the hypothesis that an increase low-risk stimulation may be beneficial for increased seizure reduction.

### Stronger circadian rhythms correspond to improved patient outcomes

Epilepsy patients have been shown to have multiple nested rhythms of both interictal activity and seizures (Baud *et al*., 2018). We therefore quantified circadian rhythmicity in the number of detected epileptiform events to determine if patients with improved outcomes exhibited different circadian patterns. This question is centered around the hypothesis that if epileptiform activity was clustered in a narrower window of the day, the patient would spend more time daily in low-risk states. Our patient cohort had similar clusters to those identified by Baud *et al*., 2018, with the majority experiencing nightly peaks in detections, specifically between 5 PM to 9 AM (Fig 4A). Dividing patients in three 8-hour partitions, we found that 13 patients had a late evening clustering pattern (5 PM – 1AM), 20 had an early morning clustering (1 AM – 9 AM), and 7 patients had an afternoon clustering (9 AM – 5 PM). Out of 40 patients, 13 patients had phase-locking values less than 0.5, which indicated weaker circadian patterns in their detections. The strength of the circadian phase-locking of epileptiform activity correlated with seizure reduction (Fig 4B; Pearson’s correlation; *p* = 0.045) and may also explain why patients with improved responses spend more time in low-risk states daily.

**Fig. 4.**
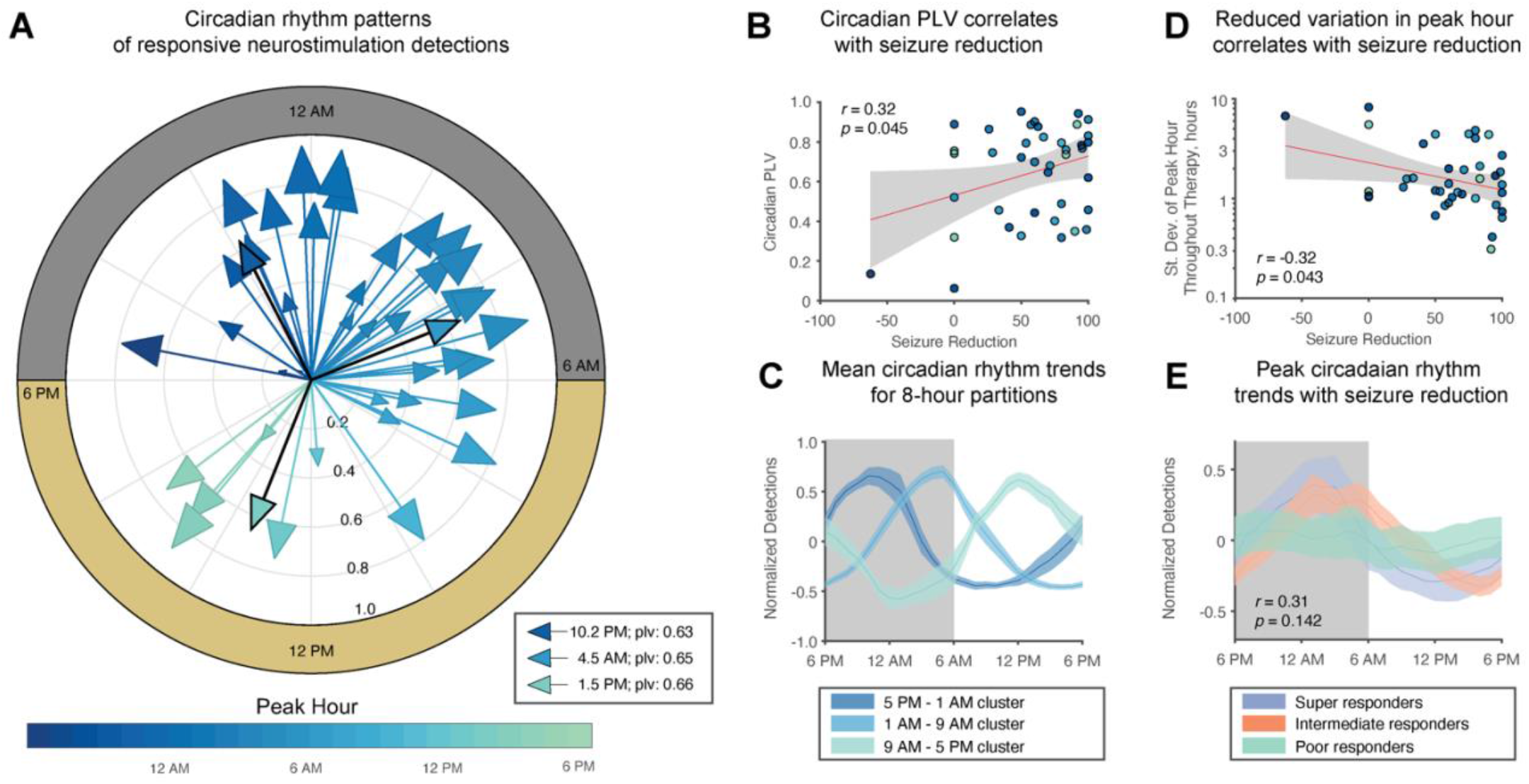
Stronger circadian rhythms of detections correspond to improved patient outcomes. (**A**) Most patients had circadian patterns in detections that peak in the night time. In three, 8-hour partitions, 13 patients had a late evening clustering pattern (5 PM – 1AM) with a peak hour at 10.2 PM, 20 patients had an early morning clustering (1 AM – 9 AM) with a peak hour at 4.5 AM, and 7 patients had an afternoon clustering (9 AM – 5 PM) with a peak hour at 1.5 PM. A larger phase-locking value (PLV) signifies a greater extent detections were concentrated to a certain hour of the day. Patients with a larger PLV experienced improved seizure reduction (Pearson’s correlation; *p* = 0.045). (**C**) Individual traces of 8-hour partition groups—in high agreement with prior literature (Baud *et al*., 2018). (**D**) A more stable peak hour throughout therapy was correlated to seizure reduction (Pearson’s correlation; *p* = 0.043). (**E**) The patient cohort exhibits a trend that patients with circadian rhythms with peaks shortly after midnight had improved outcomes (Circular-linear correlation; *p* = 0.142). The weight peak hour was 0.8 AM for super responders, 3.3 AM for intermediate responders, and 22.7 PM for poor responders.

Additionally, we found that a more stable peak hour was correlated to seizure reduction (Pearson’s correlation; *p* = 0.043) when we quantified the standard deviation of circadian peak hour for each patient in 45-day intervals throughout therapy (Fig 4D). In the 8-hour clusters, groups had average peak hours at 10.2 PM, 4.5 AM, and 1.5 PM (Fig 4C) for late evening, early morning, and afternoon clusters, respectively. Finally, we observed an insignificant trend that patients with circadian rhythms peaking shortly after midnight had improved outcomes (Fig 4E; circular-linear correlation; *p* = 0.142).

### Phase-locking of long episodes to patient rhythms

We additionally quantified the extent that long episodes, or periods of repeated detections which can be used as a surrogate for seizures (Fig 2C), were phase-locked to each patient’s unique circadian cycle and most prominent multidien cycle. In high agreement with Baud *et al*., 2018, we found that the majority of patients received stimulation between the rising phase and peak phase of daily detections (Fig 5A). Similarly, for peak multidien rhythms, long episodes occurred between the rising and peak phases of the multidien rhythm (Fig 5B). There was no correlation between the phase angle of long episodes on circadian (circular-linear correlation; *p* = 0.91) or multidien (circular-linear correlation; *p* = 0.90) cycles and seizure reduction.

**Fig. 5.**
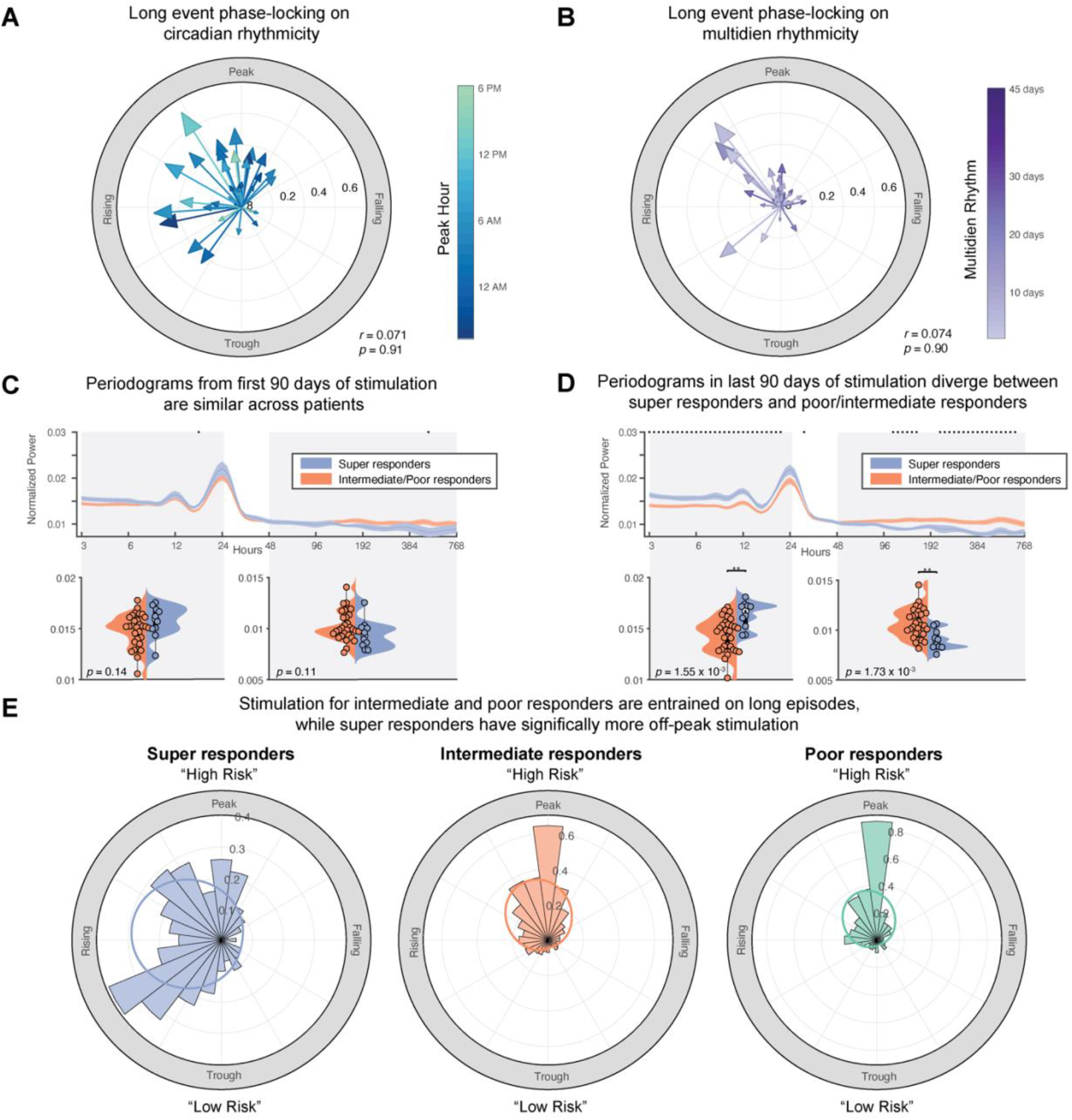
Periodograms of super responders and poor/intermediate responders diverge over time, and stimulation of super responders is less phase-locked to long episodes. (**A**) We found that most patients received stimulation between the rising phase and peak phase of daily detections. Seizure reduction did not significantly correlate with phase angle of long episodes on the circadian cycle (Circular-linear correlation; *p* = 0.91) (**B**) For peak multidien rhythms, long episodes occurred between the rising and peak phase of the most prominent multidien rhythm (≥ 48 hours). Seizure reduction did not significantly correlate with phase angle of long episodes on the circadian cycle (Circular-linear correlation; *p* = 0.90). Periodograms are not significantly different at the start of stimulation therapy between super responders and intermediate/poor responders. (**D**) In the last 90 days of stimulation, super responders experienced increased power in short periods (≤ 24 hours) (two-sample *t*-test, *p* = 1.55 × 10^−3^) while intermediate/poor responders had increased power in multidien periodics (≥ 48 hours) (two-sample *t*-test, *p* = 1.73 × 10^−3^). (**E**) Across all periods from 3 hours to 45 days, there was a high level of phase-locking between long episodes and stimulation episodes in intermediate and poor responders. However, the distribution of stimulation episodes and long episodes between super responders and both intermediate (2-sample Kuiper test; *p* < 0.01) and poor responders (2-sample Kuiper test; *p* < 0.01) were significantly different, indicating more stimulation occurred outside of high-risk long episodes in super responders. Von mises distributions for super responders (µ: -1.39 rad, κ = 0.74), for intermediate responders (µ: -0.36 rad, κ = 1.06), and for poor responders (µ: -0.36 rad, κ = 1.15).

To quantify how rhythms of epileptiform activity evolved over the course of stimulation, we calculated patient periodograms across periods spanning 3 hours to 32 days from the first 90 days of stimulation therapy to the last 90 days of stimulation. Super responder and intermediate/poor responder patients diverged significantly in the last 90 days of stimulation. Specifically, super responder patients had increased power in short periods (≤ 24 hours; Fig 5 C,D; two-sample *t*-test, *p* = 1.55 × 10^−3^) while intermediate/poor responders had increased power in multidien periodics (≥ 48 hours; Fig 5 C,D; two-sample *t*-test, *p* = 1.73 × 10^−3^). We suspect a reduction in power during multidien periods, as observed in the super responder group, indicates that seizures reoccurring on slower rhythms become less prominent and the respective increase in power in short periodics (≤ 24 hours) may indicate a restoration of normal circadian rhythms.

We finally quantified the phase angle distributions for long episodes and stimulation episodes across the entire therapy duration for periods ranging from 3 hours to 45 days (Fig 5E). A greater concentration of bins at the peak indicates that most stimulation occurs in phase with long episodes, i.e. seizures. For intermediate and poor responders, there was a high level of phase-locking between long episodes and stimulation, as one would expect given the goal of closed-loop stimulation in response to epileptiform activity (Fig 5E). Interestingly, the distribution for super responder patients was significantly different compared to both intermediate (2-sample Kuiper test; *p* < 0.01) and poor responders (2-sample Kuiper test; *p* < 0.01). This demonstrates that super responders had increased instances of stimulation outside of the peak of long episode events compared to intermediate and poor responders.

## DISCUSSION

In this work, we present evidence that stimulation during brain states defined by higher or lower risk for seizures may be critical to improving patient outcomes with responsive neuromodulation for epilepsy. Our results contain several notable findings: 1) the amount of stimulation and time in low-risk states early in therapy is a predictor of outcomes, which increases steadily over time and matches reports of long-term improvements, 2) super responders had more narrow and consistent daily clustering of epileptiform activity than poor and intermediate responders, serving as an additional prognostic factor, and 3) patients with improved outcomes had more stimulation episodes out of phase with long episodes, i.e. periods of repeated detections correlated with seizures. Together, these findings suggest that the presence of, and stimulation during, low-risk brain states both favor improved outcomes with closed-loop responsive neurostimulation.

State-dependent neurostimulation has been actively investigated for several neurological disorders, such as Parkinson’s disease (Gilron *et al*., 2021; Louie *et al*., 2021) and Tourette’s syndrome (Cagle *et al*., 2022). In the context of epilepsy, closed-loop intracranial neurostimulation targets epileptiform activity. While sometimes conceived as only targeting seizures for stimulation, most RNS System stimulation events happen in response to interictal epileptiform activity, hundreds to thousands of times daily, rather than seizures themselves, which occur at far lower rates. This makes the ratio of stimulation in the interictal vs. ictal period based on baseline seizure rate more than 1000:1 for most patients (Fig S2). As we hypothesize in Figure 1C, perhaps much of the functional changes observed in patients who respond to RNS System therapy are due to stimulation during “low-risk” interictal states, rather than higher risk peri-ictal states (Chiang *et al*., 2021; Khambhati *et al*., 2021).

To understand the motivation for this hypothesis, we return to basic principles of plasticity of neural connections. Seizures themselves are events that can facilitate future seizures through strengthening of excitatory connections (Morimoto, Fahnestock and Racine, 2004), and brain networks may undergo reorganization due to excess excitation (Bains, Longacher and Staley, 1999; Jarero-Basulto *et al*., 2018). While the role of neuroplasticity is critical in the generation of epilepsy, harnessing neuroplasticity may be similarly important for neuromodulation approaches to alter epileptic brain networks to restore healthy function. It has been reported that tissue taken from epileptic rodent models and human epilepsy patients exhibits impaired synaptic plasticity (Beck *et al*., 1996; Abegg *et al*., 2004). In human patients implanted with intracranial electrodes, memory tasks – which rely on plasticity – are impaired in the presence of interictal spiking (Liu and Parvizi, 2019; Reed *et al*., 2020; Leeman-Markowski *et al*., 2021; Camarillo-Rodriguez *et al*., 2022). Taking this information into account, it seems plausible that the most opportune time to elicit lasting change to network connections that might prevent seizures from recurring in the future might be during low-risk epochs when learning is more likely to occur and be maintained.

We defined the time spent in, and stimulation during, low-risk periods, specifically demarcated by days with fewer or equal numbers of repetitive detections known as “long episodes” when compared to the average detection counts from the prior week. We found that quantification of low-risk time and stimulation episodes were predictive of seizure reduction within the first 90 days of stimulation (Fig 3A, B). Further, patients with a greater excess of high-risk stimulation episodes per seizure than chance exhibited worse outcomes (Fig 3E), an indication that low-risk stimulation likely plays an important role in therapy in addition to high-risk stimulation.

Additional evidence that stimulation during periods of reduced epileptiform activity may be important for inducing long-term changes in brain dynamics that suppress seizures is the comparable success seen in open-loop deep brain stimulation for epilepsy. While there has not been a clinical trial directly comparing the outcomes of deep brain stimulation (DBS) for epilepsy and responsive neurostimulation, the outcomes from both groups are relatively comparable (Klinger and Mittal, 2018). According to a systematic review of hippocampal deep brain stimulation, a close parallel to the frequent mesial temporal locations of responsive stimulation leads, the mean seizure reduction was 67.8% (Vetkas *et al*., 2022). For generalized epilepsies using DBS, anterior thalamic nucleus and centromedian thalamic nucleus lead locations had a 60.8% and 73.4% mean seizure reduction (Vetkas *et al*., 2022). In a comparison of patients with temporal lobe epilepsy with DBS and the RNS System, median seizure reduction was 58% after 12-15 months for 26 ANT-DBS patients and 70% for 32 patients with TL-RNS (Yang *et al*., 2022). DBS for epilepsy does not produce a continuous train of pulses as typical for movement disorders, but rather follows a 1-minute ON cycle and 5-minute OFF cycle. Given that stimulation timing is non-specific to seizure state in DBS, stimulation during low-risk states is guaranteed (represented by the blue line in Fig 3D).

Recent work has shown that patients experience multiscale seizure cycles that may be circadian or multidien (Baud *et al*., 2018; Gregg *et al*., 2021; Leguia *et al*., 2021). We were able to reproduce findings from Baud *et al*., 2018 and saw similar late evening, early morning, and afternoon peaks in detections on a circadian cycle in our patient cohort (Fig 4 A, C). Moreover, we found in Figure 4B that a larger circadian phase-locking value was correlated with improved seizure reduction, which may enable more “low-risk” periods daily due to stronger clustering of detections in smaller windows of the day.

We showed that the periodograms between super-responders and intermediate/poor responders diverged over the course of therapy (Fig 5 C, D), with super-responder patients demonstrating increased strength in circadian or faster rhythms compared to intermediate/poor-responders, who had stronger multidien, slower rhythms. This shift may serve as preliminary evidence that detections in super-responder patients may exhibit daily rhythms that more closely resemble rhythms one might expect in healthy subjects with no history of seizures. Finally, we observed significant differences in the distributions of phase of long episodes and stimulation episodes between super responders and poor/intermediate responders (Fig 5E). Specifically, across all periods spanning 3 hours to 45 days, intermediate and poor responders had stimulation highly entrained to long episodes. In contrast, super responders experienced more instances of stimulation out of phase with long episodes, which supported our hypothesis that low-risk stimulation may play a contributing role in patient improvement through neuromodulation therapy.

### Future Directions

Based on this evidence, it appears that the amount of time in low-risk states may be an early predictor of therapeutic outcome. While the calculations of low-risk state are related to counted detections saved from the NeuroPace RNS System, it may be possible to determine low-risk time prior to the implant of the device using intracranial EEG. This may be done using similar detection settings to the device’s line length detector or band pass range thresholding and similarly defining long episodes from prolonged detections. While our paper focused on the timing of stimulation, previous studies from our group found that patient-specific structural connections are predictive of clinical outcomes (Charlebois *et al*., 2022). It may be possible to combine both the structural and temporal aspects of stimulation for improved predictability. The amount of time spent in low-risk states could also potentially be used as a surrogate of patient improvement where reporting of seizure frequency can be imperfect. As previously reported, quantified time spent in low-risk states in Figure 3C steadily increased over time in intermediate/poor responders, which parallels the slow but steady improvement over time reported in RNS System patients in long term follow-up studies (Nair *et al*., 2020). Further, perhaps parameters of closed-loop devices could be adjusted to increase targeting of low-risk states to facilitate network changes. Additionally, if improvements in neuromodulation therapy over time are due to functional changes driven by neuromodulation-induced plasticity, stimulation may be programmed to facilitate these long-term improvements to occur earlier.

### Limitations

In this paper, we put forward a potential mechanism for successful responsive neurostimulation for epilepsy with an emphasis on neuromodulation-induced plasticity being responsible for driving network changes necessary to prevent future seizures. We were limited in this study by using retrospective data, and this study does not show causation. Patients who had better outcomes may simply be in more low-risk epochs because they have fewer seizures early on, perhaps due to a more pronounced micro-lesion effect following implantation (Oommen, Morrell and Fisher, 2005). Despite this, a divergence in low-risk time between super responder and intermediate/poor responders occurred early in therapy (first 90 days) and this metric may be useful in future work to objectively quantify improvement over time. Another limitation is the lack of full characterization of detected events by the closed-loop system. Whether detections are interictal discharges, trains of discharges, or spurious detections is not adjudicated at the individual event level. Despite this, we found correlation between baseline long episode counts and seizures, and papers using similar methodology have found similar agreement with detections of interictal epileptiform activity such as spike-waves, rhythmic alpha/beta, and low voltage fast activity. Yet, these trends remain predictive, regardless of their precise characterization. Finally, patients in this cohort remained on anti-epileptic medications concurrent with the RNS System, and we did not study whether adherence to medication coupled with the RNS System could lead to better outcomes. It is also feasible that certain drugs may put patients in low-risk periods more often, and thus facilitate neuromodulation-induced plasticity. Despite these limitations, we believe our results support further exploration of stimulation during low-risk states as a means to generate long-term functional changes seen in patients with excellent outcomes from neuromodulation.

## METHODS

### Study Design

This study aimed to quantify stimulation patterns and the temporal dynamics of low- and high-risk brain states in a cohort of 40 patients at the University of Utah who were treated with the NeuroPace responsive neurostimulation system (RNS System). All cases were performed at the University of Utah hospital from 2015-2021, and a retrospective analysis of this cohort was approved by the University of Utah Institutional Review Board. Patients were included in this study if stimulation was enabled and had a baseline seizure frequency and follow-up seizure frequency reported by a board-certified epileptologist at last follow up at least 180 days after stimulation had been enabled. Patients with psychogenic seizures (n=2) were excluded in this study to ensure accurate seizure reporting.

### Measuring clinical outcome

All clinical outcomes and monthly seizure frequency rates were recorded by board-certified epileptologists during clinical interviews. Seizure reduction was quantified as a percentage, calculated by subtracting the follow up monthly seizure frequency from the monthly baseline seizure frequency, divided by baseline seizure frequency (Eq. 1).

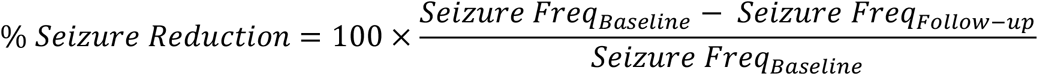

A 100% seizure reduction value indicates seizure freedom at follow-up, while a 0% seizure reduction indicates no change from baseline at follow-up. A negative seizure reduction value indicates worsening seizure frequency and only occurred for 1 patient in this cohort. Patients were categorized into three groups based on their seizure reduction values: super responders (>90% reduction, n=10), intermediate responders (≥ 50% reduction and ≤ 90% reduction), n=19, and poor responders (<50% reduction, n=11) (Figure 1A). In analyses where outcomes were not analyzed on a continuum, intermediate and poor responders were grouped together (n=30) to be compared to super responders (n=10). This grouping was done to capture all potential patient variability and identify major differences from patients with subpar outcomes to those with ideal outcomes from therapy.

### Calculation of daily risk periods

Calculation of low-risk periods was motivated by prior work from Chiang *et al*., 2021, who defined optimal stimulation parameters based on high- and low-risk states using histogram data extracted from the RNS System. We used long episode counts and saturations as exported from the RNS System to determine risk states in this study. Depicted in Figure 1D, daily long episodes counts were retrieved and convolved with a normalized kernel of [1/4, 1/2, 1/4] days to ensure that counts from the previous day and following day factored into risk assessment for each day. To determine whether a period is high risk, the average number of convolved long episodes from the previous week is used as a threshold. If the daily risk amplitude is greater than the prior 7-day threshold, then the day is considered high-risk, while if the value is less than or equal to the prior 7-day threshold, then the day is considered low-risk. We used a 7-day window to define a moving risk metric rather than taking the average over the entire time series to account for instances when programming settings were changed, potentially altering the number of long episodes detected. For patients who did not reliably sync their device and have gaps in their histogram data, we followed established methods (Baud *et al*., 2018; Gregg *et al*., 2021) to fill in the gaps in the histogram data when fewer than 12 hours of histogram data were logged in one day. An example of this method can be found in Supplementary Figure S3.

### Circadian and multidien rhythms

Circadian and multidien rhythms were calculated from the patient histogram data using number of detections across all RNS channels following similar methods as described in (Baud *et al*., 2018; Gregg *et al*., 2021; Leguia *et al*., 2021). The time-series data was z-scored within each programming epoch to accommodate changes to detection settings during programming sessions. Spectrograms were produced for each patient by a continuous wavelet transform (Morlet) on the z-scored detection counts from a periodic range of 3 hours to 45 days over 91 periodic bins. The CircStat toolbox was used to calculate mean angle and mean resultant vector of the peak hour on a 24-hour cycle. To determine phase locking, the mean angle and mean resultant vector was calculated by measuring the entrainment of long episodes on either the circadian or largest prominent multidien peak (≥ 48 hours). Patients with long episodes occurring slower than 45-day periods (n=2) were excluded from the phase-locking analysis. The first and last 90-day window since stimulation was used to calculate an average periodogram for each patient. The 90-day periodogram was calculated by averaging each periodic time band from the spectrogram produced by a continuous wavelet transform (Morlet) on the z-scored detection counts from a periodic range of 3 hours to 32 days. The instantaneous phase and power were calculated over 81 periodic bins, and the spectrogram was L1 normalized to avoid the attenuation of amplitude at shorter periodic cycles. All periodograms per patient were confirmed to sum up to 1. In order to compare changes in periodicity over time, the first 90 days of stimulation were compared to periodograms generated in the 90 days before the follow-up outcome collected. To quantify phase-locking of long episodes to stimulation, we repeated the analysis used with detections counts using stimulation episode counts instead. We plotted circular histograms of the probability distribution function of mean angles of long episodes and stimulation episodes for all periodics ranging from 3 hours to 45 days. Von Mises distributions were calculated using the CircStat toolbox for each responder group and visualized over top the circular histogram plots.

### Statistics

All analyses and statistics were completed in MATLAB 2021b. Unpaired statistical tests used a Student’s t-test, and paired statistics tests used two-sample t-tests. For data following a circular distribution, we used the CircStat toolbox, a circular statistics toolbox written for MATLAB (Berens, 2009), to determine correlation and significance. Additionally, circular 2-sample Kuiper tests were used from the CircStat toolbox to test whether phase-locking distributions were from the same distribution. All shading in plots are defined by the mean ± standard error.

## Data Availability

All de-identified data are available upon request to the corresponding author.

## List of Supplementary Materials

Fig S1. Epilepsy duration, patient age, and age of epilepsy onset do not correlate with outcome.

Fig S2. Daily stimulation episodes far exceed daily seizures.

Fig S3. Addressing data gaps with interpolation and variability in detection counts with z-scoring.

## Author contributions

Conceptualization: DNA, JDR

Methodology: DNA, CMC, EHS, AMA, BJN, AYP

Data Curation: DNA, CMC, TSD, CRB, JDR Visualization: DNA

Funding acquisition: DNA, JDR

Project administration: JDR

Supervision: JDR, KSW

Writing – original draft: DNA

Writing – review & editing: JDR, TSD, EHS, CMC, KSW

## Data and materials availability

All de-identified data are available upon request to the corresponding author.

## References and Notes

Abegg, M. H. et al. (2004) ‘Epileptiform activity in rat hippocampus strengthens excitatory synapses’, Journal of Physiology, 554(2), pp. 439–448. doi: 10.1113/jphysiol.2003.052662.

Bains, J. S., Longacher, J. M. and Staley, K. J. (1999) ‘Reciprocal interactions between CA3 network activity and strength of recurrent collateral synapses’, Nature Neuroscience, 2(8), pp. 720–726. doi: 10.1038/11184.

Baud, M. O. et al. (2018) ‘Multi-day rhythms modulate seizure risk in epilepsy’, Nature Communications. Nature Publishing Group, 9(1). doi: 10.1038/S41467-017-02577-Y.

Beck, H. et al. (1996) ‘Synaptic Plasticity in the Human Dentate Gyrus’, Thus, The Journal of Neuroscience, 20(18), pp. 7080–7086.

Berens, P. (2009) ‘CircStat: A MATLAB Toolbox for Circular Statistics’, Journal of Statistical Software, 31(10), pp. 1–21.

Bergey, G. K. et al. (2015) ‘Long-term treatment with responsive brain stimulation in adults with refractory partial seizures’. Available at: https://www.clinicaltrials.gov x(Accessed: 26 April 2022).

Cagle, J. N. et al. (2022) ‘for Tourette Syndrome’, 32611, pp. 1–5. doi: 10.1001/jamaneurol.2022.2741.

Camarillo-Rodriguez, L. et al. (2022) ‘Temporal lobe interictal spikes disrupt encoding and retrieval of verbal memory: A subregion analysis’, Epilepsia, 63(9), pp. 2325–2337. doi: 10.1111/epi.17334.

Charlebois, C. M. et al. (2022) ‘Patient-specific structural connectivity informs outcomes of responsive neurostimulation for temporal lobe epilepsy’, Epilepsia, 63(8), pp. 2037–2055. doi: 10.1111/epi.17298.

Chiang, S. et al. (2021) ‘Evidence of state-dependence in the effectiveness of responsive neurostimulation for seizure modulation’, Brain Stimulation. Elsevier, 14(2), pp. 366–375. doi: 10.1016/J.BRS.2021.01.023.

Gilron, R. et al. (2021) ‘Long-term wireless streaming of neural recordings for circuit discovery and adaptive stimulation in individuals with Parkinson’s disease’, Nature Biotechnology. Springer US, 39(9), pp. 1078–1085. doi: 10.1038/s41587-021-00897-5.

Gregg, N. M. et al. (2021) ‘Thalamic deep brain stimulation modulates cycles of seizure risk in epilepsy’, Scientific Reports. Nature Publishing Group UK, 11(1), pp. 1–12. doi: 10.1038/s41598-021-03555-7.

Jarero-Basulto, J. J. et al. (2018) ‘Interactions between epilepsy and plasticity’, Pharmaceuticals, 11(1), pp. 1–18. doi: 10.3390/ph11010017.

Khambhati, A. N. et al. (2021) ‘Long-term brain network reorganization predicts responsive neurostimulation outcomes for focal epilepsy’, Science Translational Medicine, 13(608), p. eabf6588. doi: 10.1126/scitranslmed.abf6588.

Klinger, N. and Mittal, S. (2018) ‘Deep brain stimulation for seizure control in drug-resistant epilepsy’, Neurosurgical Focus, 45(2). doi: 10.3171/2018.4.FOCUS1872.

Kokkinos, V. et al. (2019) ‘Association of Closed-Loop Brain Stimulation Neurophysiological Features with Seizure Control among Patients with Focal Epilepsy’, JAMA Neurology, 76(7), pp. 800–808. doi: 10.1001/jamaneurol.2019.0658.

Leeman-Markowski, B. et al. (2021) ‘Effects of hippocampal interictal discharge timing, duration, and spatial extent on list learning’, Epilepsy and Behavior. Elsevier Inc., 123, p. 108209. doi: 10.1016/j.yebeh.2021.108209.

Leguia, M. G. et al. (2021) ‘Seizure Cycles in Focal Epilepsy Supplemental content CME Quiz at jamacmelookup.com’, JAMA Neurol, 78(4), pp. 454–463. doi: 10.1001/jamaneurol.2020.5370.

Lesser, R. P. et al. (1999) ‘Brief bursts of pulse stimulation terminate afterdischarges caused by cortical stimulation’, Neurology, 53(9), pp. 2073–2081. doi: 10.1212/wnl.53.9.2073.

Liu, S. and Parvizi, J. (2019) ‘Cognitive refractory state caused by spontaneous epileptic high-frequency oscillations in the human brain’, Science Translational Medicine, 11(514), pp. 1–14. doi: 10.1126/scitranslmed.aax7830.

Louie, K. H. et al. (2021) ‘Gait phase triggered deep brain stimulation in Parkinson’s disease’, Brain Stimulation, 14(2), pp. 420–422. doi: 10.1016/j.brs.2021.02.009.

Morimoto, K., Fahnestock, M. and Racine, R. J. (2004) ‘Kindling and status epilepticus models of epilepsy: Rewiring the brain’, Progress in Neurobiology, 73(1), pp. 1–60. doi: 10.1016/J.PNEUROBIO.2004.03.009.

Morrell, M. J. (2011) ‘Responsive cortical stimulation for the treatment of medically intractable partial epilepsy’, Neurology, 77(13), pp. 1295–1304. doi: 10.1212/WNL.0b013e3182302056.

Naik, A. A. et al. (2021) ‘Mechanism of seizure-induced retrograde amnesia’, Progress in Neurobiology. Elsevier Ltd, 200(July 2020), p. 101984. doi: 10.1016/j.pneurobio.2020.101984.

Nair, D. R. et al. (2020) ‘Nine-year prospective efficacy and safety of brain-responsive neurostimulation for focal epilepsy’, Neurology. Mayo Clinic Minnesota, 95(9), pp. e1244–e1256. doi: 10.1212/WNL.0000000000010154.

Oommen, J., Morrell, M. J. and Fisher, R. (2005) ‘Experimental Electrical Stimulation Therapy for Epilepsy’, Current Treatment Options in Neurology, 7(4), pp. 261–271. doi: 10.1891/9781617051876.0030.

Reed, C. M. et al. (2020) ‘Extent of single-neuron activity modulation by hippocampal interictal discharges predicts declarative memory disruption in humans’, Journal of Neuroscience, 40(3), pp. 682–693. doi: 10.1523/JNEUROSCI.1380-19.2019.

Rønborg, S. N. et al. (2021) ‘Acute effects of brain-responsive neurostimulation in drug-resistant partial onset epilepsy’, Clinical Neurophysiology. Elsevier Ireland Ltd, 132(6), pp. 1209–1220. doi: 10.1016/J.CLINPH.2021.03.013.

Vetkas, A. et al. (2022) ‘Deep brain stimulation targets in epilepsy: Systematic review and meta-analysis of anterior and centromedian thalamic nuclei and hippocampus’, Epilepsia, 63(3), pp. 513–524. doi: 10.1111/epi.17157.

Yang, J. C. et al. (2022) ‘Anterior nucleus of the thalamus deep brain stimulation vs temporal lobe responsive neurostimulation for temporal lobe epilepsy’, Epilepsia. doi: 10.1111/epi.17331.

