## Supplemental for "Closed-loop neurostimulation for epilepsy leads to improved outcomes when stimulation episodes are delivered during periods with less epileptiform activity"

### Supplementary Figures

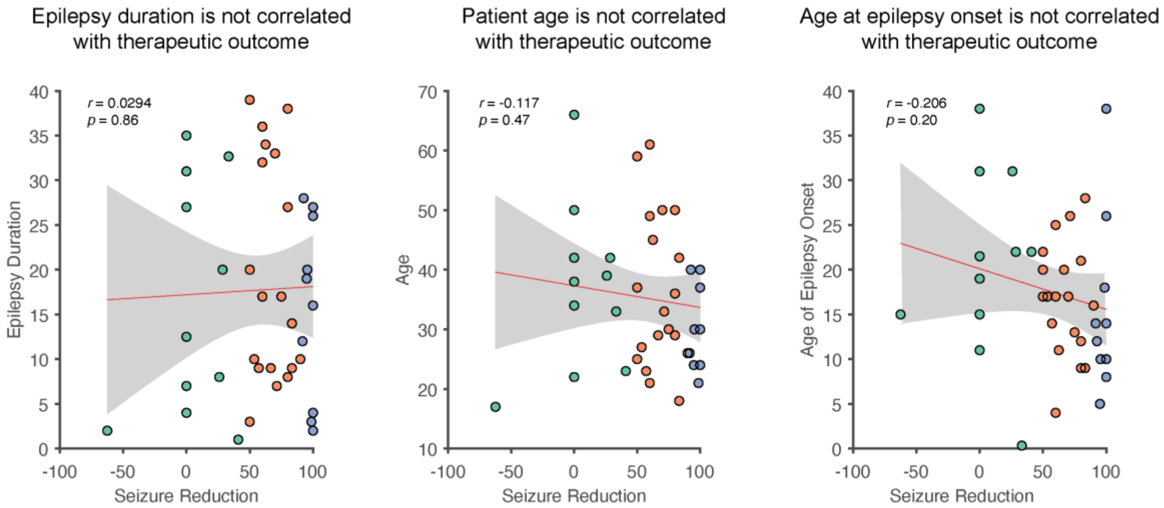


**Figure S1. Epilepsy duration, patient age, and age of epilepsy onset do not correlate with outcome.** There is no significant correlation between patient outcomes and epilepsy duration, patient age at the time of device implant, or age of epilepsy onset. Blue circles are super responders, orange circles are intermediate responders, green circles are poor responders.

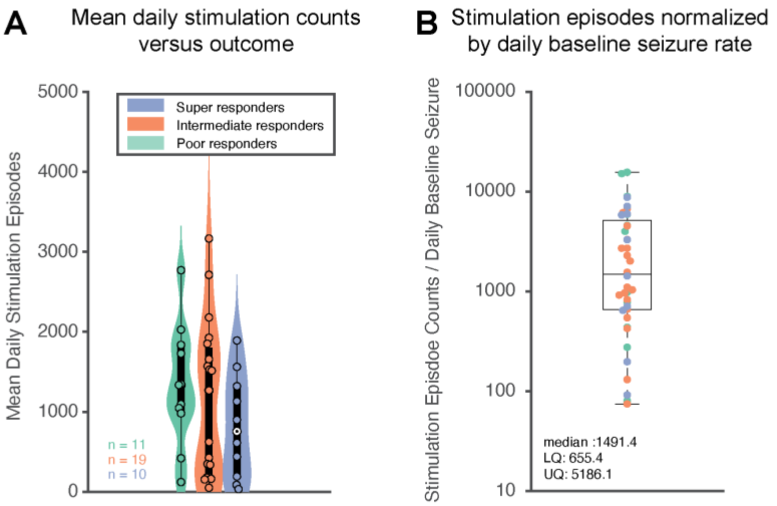


**Figure S2. Daily stimulation episodes far exceed daily seizures.** (**A**) There is no significant difference between the number of stimulation episodes across super responders, intermediate responders, and poor responders. Median daily stimulation episode counts for the duration of therapy is 1112.9 episodes/day (LQ: 341.330, UQ: 1692.987). (**B**) Average daily stimulation normalized by baseline daily seizure rate demonstrates that patients have far greater stimulation episodes than seizures, indicating that the vast majority of stimulation is not given solely in response to seizures (Median 1491.4; LQ: 655.4; UQ: 5186.1).

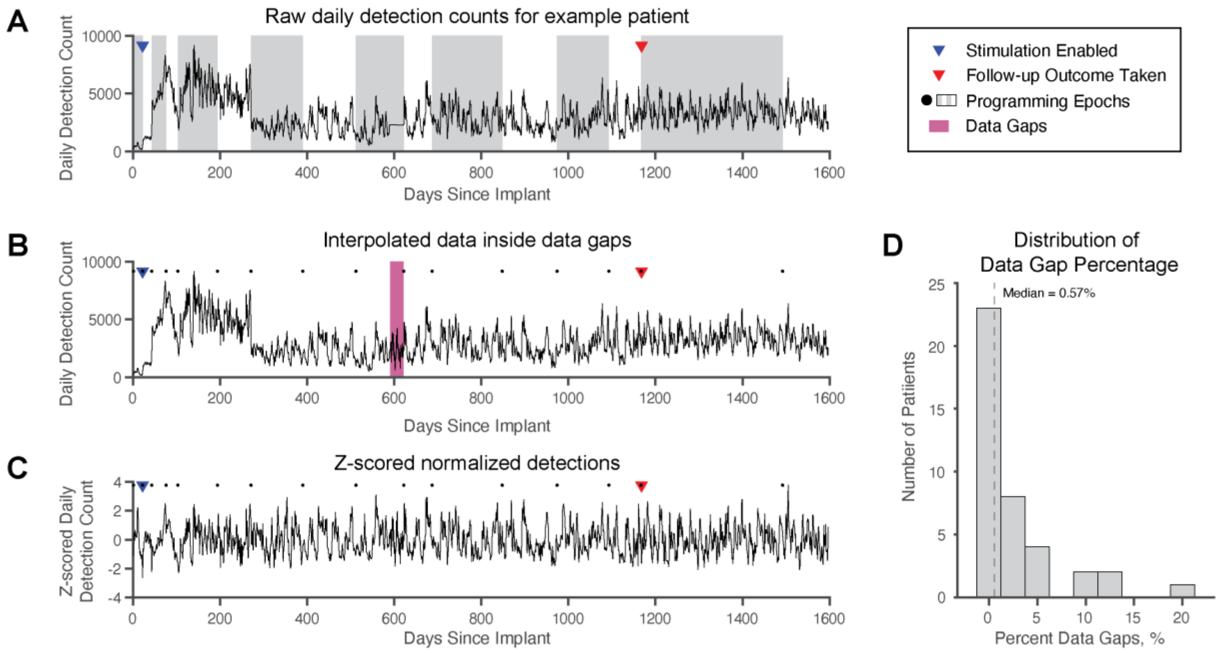


**Figure S3. Addressing data gaps with interpolation and variability in detection counts with z-scoring.** (**A**) Raw detection counts from patient histograms can be visualized over the course of therapy and correspond to interictal epileptiform discharges. Sudden changes in detection counts can occur due to changes in detection settings within each programming epochs (shaded gray area). (**B**) Patient data streams can have gaps, marked in pink, when the patient did not regularly upload data and older histogram data was overwritten to accommodate new data. Days with less than <12 hours of histogram counts were also considered to be a missing. Following methods developed by Baud et al., 2018, we interpolated data in the gap periods using known data the same length of the gap period on each side of the gap. We found a linear fit between the two ends of the gap period and applied Gaussian noise using the standard deviation of flanking regions. (**C**) When raw data is z-scored to ensure regularity in amplitudes, z-scoring occurs within programming epochs (annotated by black dots). (**D**) Most patients experience few data gaps, with a median gap percentage of 0.57% (minimum: 0%, maximum 22.5%).
